# Endoscopic Predictors of Early Cancer Burden Allow Optimization of Management Decisions in Hereditary Diffuse Gastric Cancer

**DOI:** 10.1101/2025.10.13.25337824

**Authors:** Lianlian Wu, Hui Jun Lim, Nandini Karthik, Sophie Samra, Maria O’Donovan, J Robert O’Neill, Marc Tischkowitz, Rebecca C Fitzgerald, Massimiliano Di Pietro

## Abstract

**Introduction:** Hereditary diffuse gastric cancer (HDGC) associated with *CDH1* germline pathogenic variants (GPV), carries a high lifetime risk of signet ring cell carcinoma (SRCC). Currently, there is uncertainty regarding to whom and when to recommend prophylactic total gastrectomy (PTG). We hypothesise a small number of early SRCC lesions correlates with low risk of progression to clinical gastric cancer. This study aimed to identify predictors of early SRCC burden and provide evidence to inform best surveillance intervals.

**Methods:** We analyzed data from 53 *CDH1*-GPV carriers who underwent PTG prospectively recruited between Aug 2004 and Aug 2024 (PTG dataset) and a separate cohort of 94 *CDH1 CDH1*-GPV carriers with ≥2 surveillance endoscopies (endoscopy dataset). Targeted biopsies (TB) and systematic random biopsies (RB) were obtained using the Cambridge protocol. Multivariable negative binomial regression was used to assess the association of clinical (age, family history, *CDH1* variant type) and endoscopic factors (mean number of positive TB and RB per endoscopy) with SRCC burden in PTG specimens. A logistic regression model with significant predictors was trained to classify patients into low and high SRCC burden. Temporal trends in biopsy findings were analyzed using linear mixed-effects models, and pathological outcomes at 6-, 12- and 24-month intervals were compared in endoscopy dataset.

**Results:** Of the 53 patients, 89% had early-stage cancer (pT1aN0M0) and 11% had no cancer (pT0N0M0). The number of SRCC foci ranged from 0 to 273 (median 33, IQR 2–37). The number of positive targeted (*P* = 0.003) and random biopsies (*P* < 0.001) during endoscopy surveillance were independent predictors of SRCC burden in the PTG specimen, whereas age, CDH1 mutation type (truncating vs. non-truncating), number of 1st and 2nd degree relatives (SDRs) were not significantly associated. In the endoscopy surveillance cohort, the number of positive biopsies remained largely stable over time, showing fluctuations rather than consistent progression; no significant temporal increase in biopsy positivity was detected over time (*P* = 0.177) with stable number of SRCC foci at follow-up. Extending surveillance intervals from 6 to 12 or 24 months did not significantly alter progression detection rates.

**Conclusion:** Endoscopic surveillance using targeted and random biopsies by experienced endoscopists provides a reliable estimate of SRCC burden in HDGC. Our findings suggest that extending surveillance intervals in patients with low early SRCC burden is clinically safe.

## Introduction

Hereditary diffuse gastric cancer (HDGC) is an autosomal dominant cancer syndrome linked to germline pathogenic variants (GPV) of *CDH1* gene, that predisposes to diffuse gastric and lobular breast cancer.^1^ The lifetime risk of gastric cancer is estimated to be 10-42%; these estimates have significantly declined in the last decade as germline genetic testing including multigene panels have become more widespread in turn leading to a reduction in ascertainment bias.^2,3^ Most recent guidelines recommend considering prophylactic total gastrectomy (PTG) for *CDH1*-GPV carrier to reduce the lifetime risk of cancer related death and recommend PTG when there is evidence of signet ring cell carcinoma (SRCC) on endoscopic biopsies.^4^ Total gastrectomy is a life changing operation, which carries significant impact on eating behaviour and body image perception, with 80% of patients having at least one GI symptom at 12 months from the operation.^5^ Two recent longitudinal studies have challenged the recommendation to surgery in this group of people, indicating that, while SRCC foci are seen in the vast majority of patients with HDGC, progression to advanced cancer is rare under active endoscopic surveillance. In these studies stage 2 cancer or higher occurred only in 2 out of 212 surveyed patients, who declined the offer of surgery on the basis of endoscopic findings.^6,7^ Lee et al, also provided evidence that some individuals with positive endoscopic biopsies, even in the presence of subtle endoscopically visible lesions, can be safely monitored for several years, without clinical progression of the disease.^7^ This implies that early SRCC lesions in HDGC can be indolent and do not all mirror the clinical features of early sporadic gastric cancer. However, analysis of PTG specimens shows a great range in the number of foci from none or very few to more than 200, suggesting that even in the context of T1a disease, the individual risk could be very different.^8^ Therefore, there remains uncertainty regarding suitability and timing of endoscopic monitoring, as there are no reliable methods to risk stratify *CDH1-GPV* carriers and there are limited data to inform surveillance intervals. We have previously described and validated endoscopic criteria for detection and characterization of early lesions, to provide endoscopists with a tool to improve quality of the endoscopic examination and confidence in the endoscopic diagnosis.^8^ Here we identify predictors of early SRCC burden as a proxy of disease extent and provide evidence to best tailor surveillance intervals based on clinical findings during longitudinal surveillance of patients with both positive and negative endoscopy findings.

## Method

### Cohorts and study design

In this study, two cohorts of patients with HDGC and pathogenic or likely pathogenic germline *CDH1* (GPV) variants were analysed.

#### PTG cohort

This consisted of 53 *CDH1*-GPV individuals who underwent at least one endoscopy followed by PTG at Addenbrooke’s Hospital between August 2005 and August 2024. We included endoscopic procedures with mapping biopsies according to the Cambridge protocol in the analysis.^4,7,9^ We excluded individuals or endoscopies without diagnostic biopsies prior to PTG, and patients who received partial gastrectomy before surveillance. The total number of SRCC foci was meticulously assessed to measure cancer burden across the entire gastrectomy specimen by gastrointestinal pathologists with special expertise in HDGC. The gastrectomy specimen is opened along the greater curvature in the fresh state, pinned on to cork board and formalin fixed for 24 hours. After fixation, resections margins are embedded and the specimen is split into anterior and posterior segments. Each segment is then divided into anatomical regions (cardia, fundus, transition zone, antrum and pylorus). The entire specimen is paraffin embedded with alternate sections kept for research. All accompanying lymph nodes are also embedded. This cohort is primarily used to explore the relationship between endoscopic findings and PTG. (Figure 1A)

**Figure 1.**
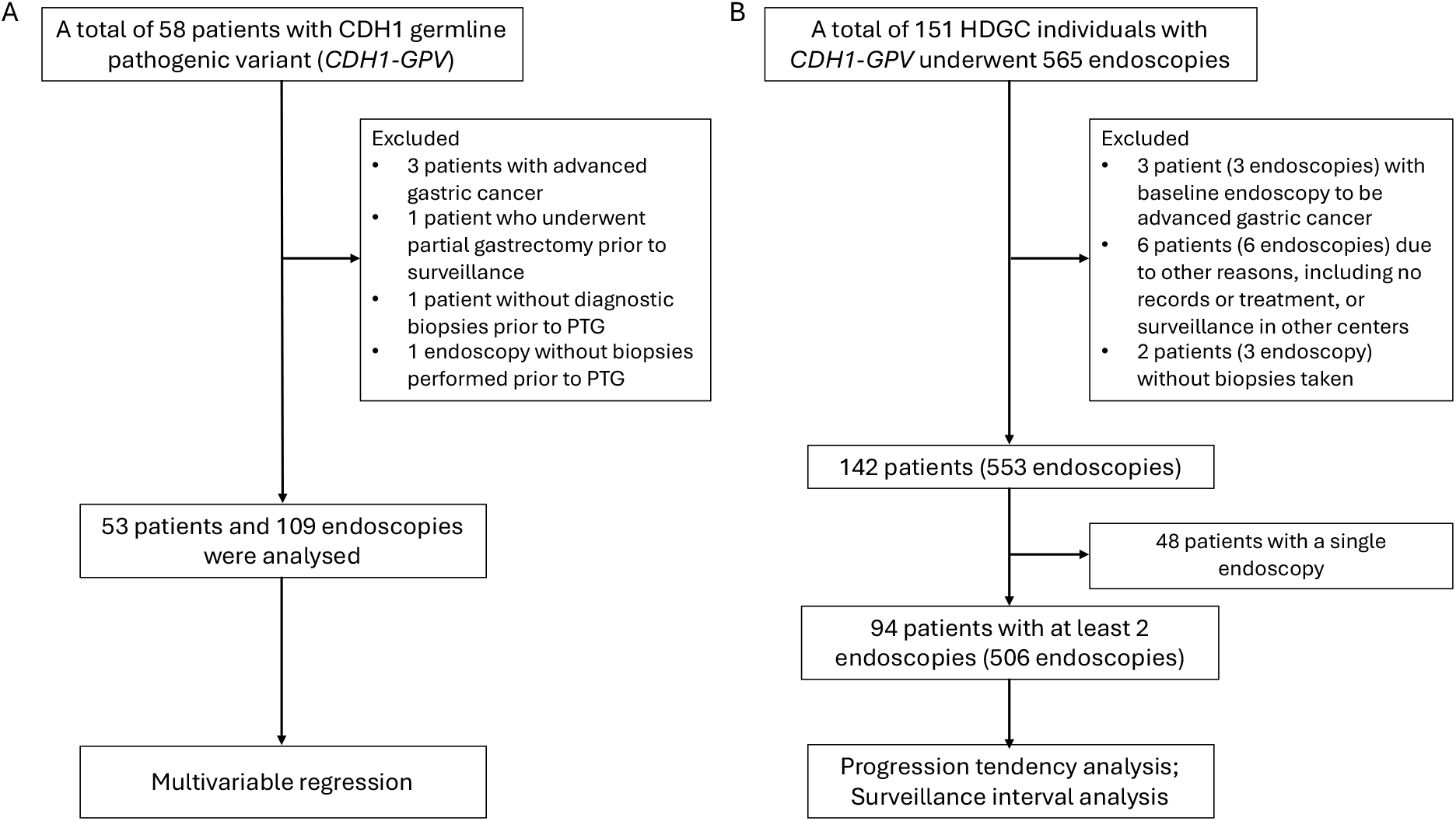
Study cohort flowcharts. (A) Flowchart outlining inclusion and exclusion criteria for the PTG (prophylactic total gastrectomy) cohort. (B) Flowchart for the endoscopy cohort. From 151 individuals who underwent 565 surveillance endoscopies, exclusions were made for negative advanced cancer at baseline, incomplete records, and single-timepoint cases, resulting in 94 individuals and 506 endoscopies.

#### Endoscopy Surveillance Cohort

This consisted of 94 *CDH1*-GPV individuals who underwent a combined total of 506 endoscopic surveillance procedures at Addenbrooke’s Hospital between June 2005 and February 2024. The exclusion criteria were as follows: baseline endoscopy revealing macroscopic evidence of invasive cancer, lack of clinical records or incomplete surveillance data, and individuals with single surveillance endoscopy (Figure 1B). This cohort is analysed for long-term disease progression of HDGC individuals under active surveillance. Patients in PTG cohort who were under surveillance are included in the endoscopy cohort.

### Procedures

Endoscopies were done with a high-resolution endoscope with optical magnification (Olympus, Tokyo, Japan). White-light imaging and narrow band imaging were used to inspect all anatomical regions of a well insufflated stomach. During endoscopies, targeted biopsies were taken from visible lesions, including pale areas, erosion, erythematous areas, ulcers or polyps. Random biopsies were performed according to the Cambridge protocol (four to five biopsy specimens in cardia, fundus, corpus, transitional zone, antrum, and pre-pyloric area, respectively) before May 1, 2019. After this date, a modified Cambridge protocol was adopted, taking two biopsies in the pre-pyloric area, four biopsies in the antrum, transitional zone, fundus, and cardia; and six biopsies in the corpus.^4,7,9^ Biopsy specimens were stained with haematoxylin and eosin (and periodic acid-Schiff diastase at discretion of the reporting pathologist) to assess for the presence of SRCC foci. All cases were reviewed by upper gastrointestinal specialist pathologists with extensive experience in the identification of SRCC.

### Multivariable regression analysis

To identify independent predictors of cancer burden in prophylactic total gastrectomy (PTG) specimens, we applied a multivariable negative binomial regression model, since the distribution of SRCC foci is skewed (Figure 2A). The response variable was the total number of signet ring cell carcinoma (SRCC) foci identified by histopathological assessment of PTG specimens. Predictor variables included patient age group at baseline, CDH1 mutation type (truncating vs. non-truncating), number of 1^st^ and 2^nd^ degree relatives (SDRs) who had symptomatic clinical gastric cancer (0, 1, or ≥2), and endoscopic findings. For endoscopic predictors, we calculated the mean number of positive targeted biopsies (TB) and random biopsies (RB) across all endoscopies prior to PTG. A second model was also fitted using only baseline endoscopy findings.

**Figure 2.**
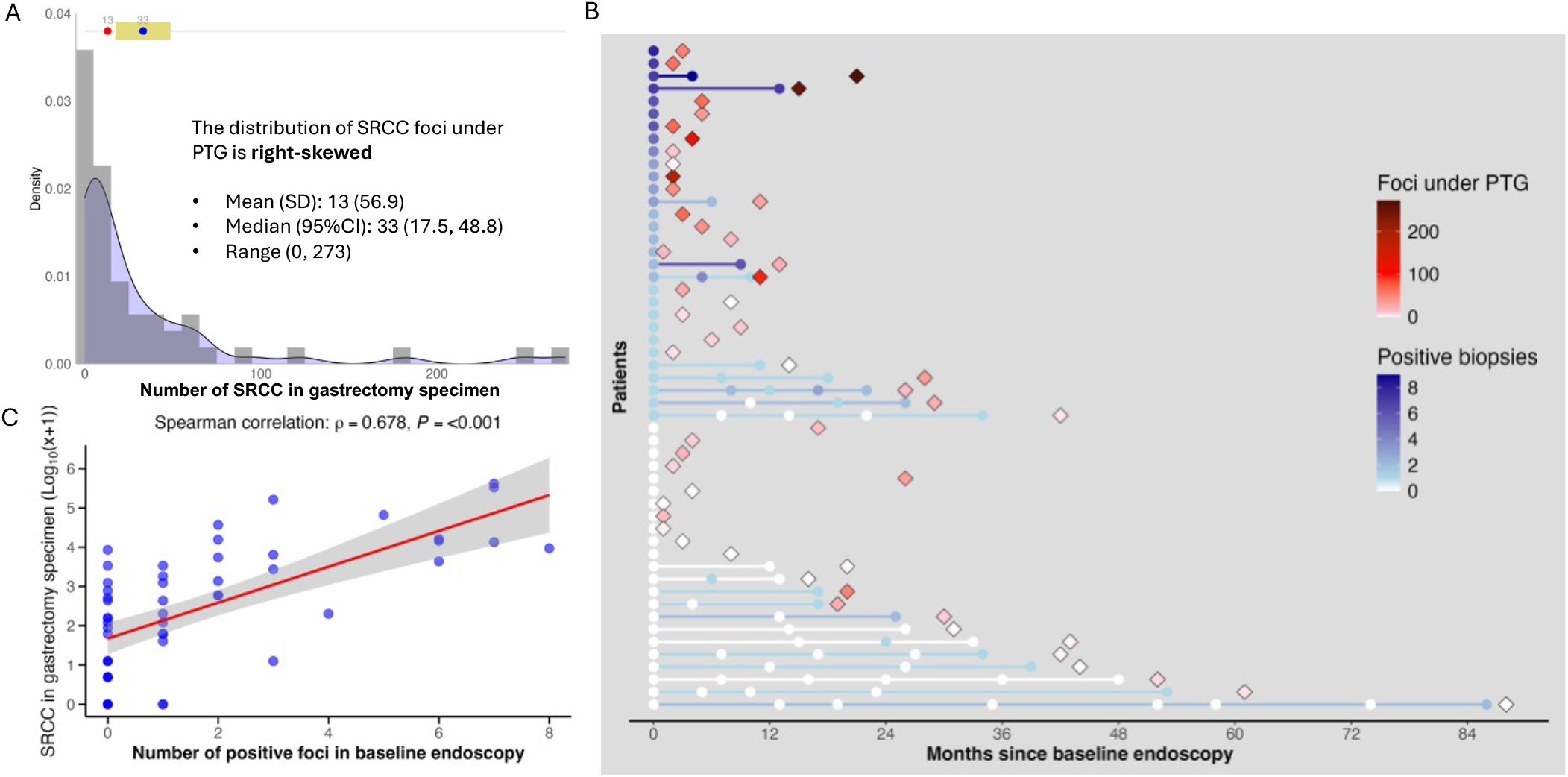
The association of endoscopic findings and cancer burden under prophylactic total gastrectomy (PTG). A. Distribution of SRCC foci in PTG specimens. Histogram showing the right-skewed distribution of SRCC foci counts among PTG patients. The majority had fewer than 50 foci, but a subset exhibited extremely high burden (>200 foci). **B. Association between endoscopic biopsy findings and signet ring cell carcinoma (SRCC) burden in gastrectomy specimens**. Each row represents one HDGC individual who underwent endoscopic surveillance followed by prophylactic total gastrectomy (PTG), and the lines were colored by the number of positive biopsies in the last endoscopy. Circles denote endoscopic procedures, colored by the number of positive biopsies, while diamonds represent PTG specimens, colored by the total number of SRCC foci identified in gastrectomy specimens. The most recent endoscopies were excluded for two patients (the second and second-to-last cases in the figure), as no biopsies were taken during their final procedure prior to PTG. A few patients without positive endoscopic biopsies underwent PTG based on their personal willingness. **C. the correlation between the number of positive foci detected at baseline endoscopy and the number of early SRCC foci identified in PTG**. Pearson correlation analysis demonstrated a moderate association, with R^2^ values of 0.445 (unadjusted) and 0.449 (adjusted for the interval between baseline endoscopy and PTG), both statistically significant (*P* < 0.001).

To evaluate the predictive utility of the endoscopic variables, logistic regression models were trained to classify patients into low (≤20 foci, n=32) or high (>30 foci, n=16) SRCC burden groups. Patients with an intermediate burden (20–30 foci, n = 5) were excluded to avoid arbitrary classification within this grey zone defined by the data distribution. Models were developed using combinations of TB and RB positivity, using either average values or baseline data. Performance was assessed using area under the receiver operating characteristic curve (AUC), sensitivity, specificity, and accuracy, averaged over 1,000 bootstrap iterations.

### Temporal Trends in Endoscopic Biopsy Findings

To investigate longitudinal changes in biopsy-detected SRCC foci, we applied mixed-effects regression models using negative binomial distribution to accommodate overdispersion in count data. The primary outcome was the number of positive foci detected per endoscopy, modelled separately for: *i*. combined random and targeted biopsies, *ii*. random biopsies only, *iii*. targeted biopsies only. The main predictor was months since baseline endoscopy, with a random intercept for each patient to account for within-subject correlation across repeated surveillance.

### Surveillance interval analysis

To evaluate the effectiveness of different surveillance intervals, we analyzed the progression status of SRCC foci in low-risk positive patients who had 1–2 positive biopsies at their first positive examination and subsequently underwent at least one further surveillance procedure. Surveillance intervals were categorized into 6-, 12-, and 24-month groups based on the time elapsed between consecutive endoscopic procedures. For the 12- and 24-month interval groups, intermediate endoscopy records (i.e., those performed within the defined interval) were masked to simulate true annual or biennial surveillance schedules. For each interval, biopsy findings were compared to those from the immediately preceding endoscopy. Progression status was defined as follows: Progressed—an increase by more than one positive biopsy; Stable—no change or an increase by only one positive biopsy; and Regressed—a decrease in the number of positive biopsies. The proportions of patients in each category were calculated and visualized to evaluate temporal patterns and the effectiveness of different surveillance intervals.

## Results

### Endoscopic surveillance findings and subsequent cancer burden under PTG

The patient characteristics of the 53 individuals who have received PTG are shown in Table 1 The cohort had a median age of 31.0 years at the time of PTG (95% CI: 27.2–34.8) and was evenly distributed by sex (49% female, 51% male). Most patients were of White ethnicity (81%), and truncating germline PV accounted for 51% of all *CDH1-PV*. The number of intramucosal SRCC foci identified in PTG specimens ranged from 0 to 273, with a median of 13 foci (Table 1).

**Table 1.** The baseline characteristics of patients underwent prophylactic total gastrectomy (n=53)

| Characteristics | Patients |
| --- | --- |
| Age at PTG, years, median (95%CI) | 31.0 (27.2–34.8) |
| Sex, Numbers (percentage) |  |
| Female | 26 (49.1%) |
| Male | 27 (50.9%) |
| Ethnicity, Numbers (percentage) |  |
| White | 43 (81.1%) |
| Asian | 5 (9.4%) |
| Others | 5 (9.4%) |
| CDH1 mutation, Numbers (percentage) |  |
| Truncating | 27 (50.9%) |
| Frameshift | 11 (20.8%) |
| Splice | 6 (11.3%) |
| Missense | 5 (9.4%) |
| Unknown | 4 (7.5%) |
| Stage, Numbers (percentage) |  |
| pT1aN0M0 | 47 (89%) |
| pT0N0M0 | 6 (11%) |
| Number of 1<sup>st</sup> or 2<sup>nd</sup> degree relatives with symptomatic gastric cancer |  |
| 0 | 16 (30.2%) |
| 1 | 15 (28.3%) |
| 2 | 17 (32.1%) |
| 3 | 4 (7.5%) |
| 4 | 1 (1.9%) |
| Number of intramucosal SRCC foci, median range) | 13 (0, 273) |

To explore the relationship between surveillance findings and cancer burden, we visualized each patient’s endoscopic results alongside the number of SRCC foci identified in their PTG specimens (Figure 2B). Patients were ordered by the number of positive biopsies detected during baseline endoscopy. This visualization revealed that individuals with higher SRCC burden were predominantly those with positive findings at baseline endoscopy identified by random and targeted biopsies. When considering only random or only targeted biopsies, the total burden could be under-estimated with more high-burden cases with a low number of SRCC lesions at endoscopy (Figure S1). Histogram showing the right-skewed distribution of SRCC foci counts from PTG specimens. The majority (81% 43/53) had no more than 50 foci, but a subset exhibited extremely high burden [4% (2/53) >200 foci] (Figure 2A). The number of positive biopsies under surveillance endoscopy showed a similar data distribution pattern (Figure S2).

To quantify this observation, we assessed the correlation between the number of positive foci detected at baseline endoscopy and the number of early SRCC foci identified in PTG. Pearson correlation analysis demonstrated a moderate association, with R^2^ values of 0.445 (unadjusted) and 0.449 (adjusted for the interval between baseline endoscopy and PTG), both statistically significant (*P* < 0.001) (Figure 2C).

### Endoscopic findings are independent predictors of cancer burden stronger than known clinical and genetic factors

To investigate the utility of endoscopic, clinical and genomic factors for SRCC-burden prediction with included into further analyses family history of cancer (SDR), age, and CDH1 mutation type (truncating vs. non-truncating). In multivariable negative binomial regression using averaged endoscopy data (Figure 3A), both the number of positive targeted biopsies (*P* = 0.003) and random biopsies (*P* < 0.001) were significantly associated with higher SRCC burden. When restricting the model to baseline endoscopy data (Figure 3B), these two predictors remained significant (*P* = 0.008 and *P* < 0.001, respectively). None of the clinical variables, including age, family history, and CDH1 mutation type, were independently associated with SRCC burden in either model.

**Figure 3.**
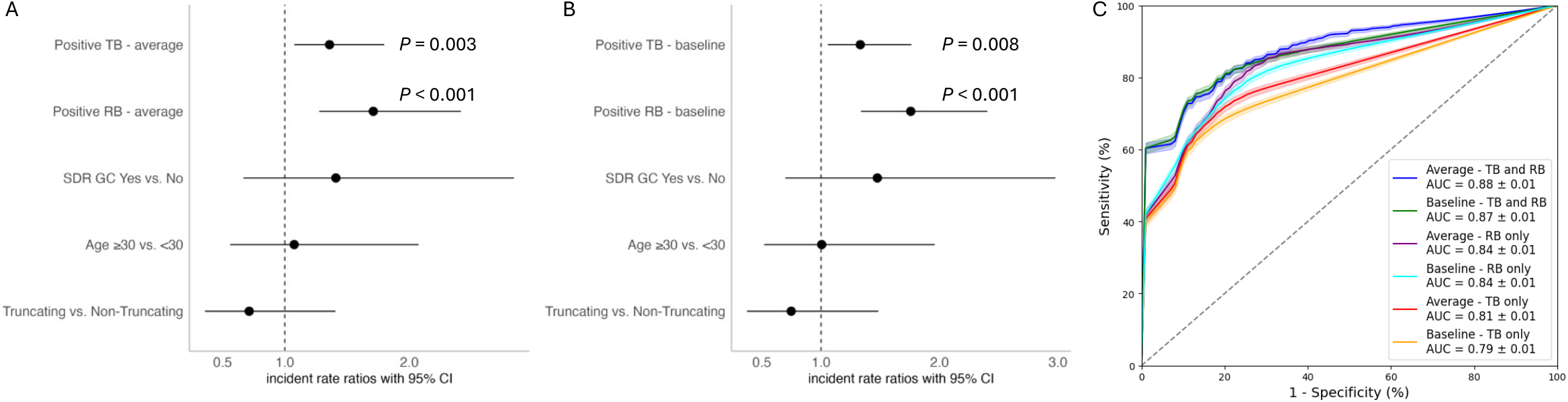
Multivariable regression model identifying independent predictors of signet ring cell carcinoma (SRCC) burden. (A) Negative binomial regression using the average number of positive targeted biopsies (TB) and random biopsies (RB) per patient showed both TB (*P* = 0.003) and RB (*P* < 0.001) were significant predictors of SRCC foci in PTG. (B) When restricted to baseline endoscopy only, TB (*P* = 0.008) and RB (*P* < 0.001) remained independent predictors. SDR: First and Secondary degree of relatives. (C) Logistic regression models trained to classify patients into high (≥30 foci) or low (≤20 foci) burden categories showed the best performance (AUC = 0.88) when both TB and RB were included.

Logistic regression models incorporating both mean TB and RB positivity achieved the best classification performance for identifying patients with high SRCC burden (Figure 3C). This model yielded an AUC of 0.88 ± 0.01. A model using only baseline TB and RB also performed well (AUC = 0.87 ± 0.01). In contrast, models using TB or RB alone showed lower performance. TB-only models had reduced sensitivity (AUC = 0.81 ± 0.01 for mean positivity, 0.79 ± 0.01 for baseline positivity), while RB-only models achieved intermediate results (AUC = 0.84 ± 0.01 for both mean and baseline positivity), underscoring the importance of combining both biopsy strategies in surveillance.

### Progression tendency analysis based on the surveillance cohort

A total of 506 endoscopic procedures from 94 *CDH1-GPV* carriers from the endoscopy cohort were analyzed to evaluate temporal trends in SRCC detection. The median follow-up time was 36 months (IQR: 19-59, maximum 159), with a median of 4 endoscopies per patient (IQR: 3-7, maximum 15). Among these individuals, 42 (45%) had no SRCC detected throughout surveillance [median follow-up 39 months (IQR: 20-57, maximum 159); median 4 endoscopies (IQR: 3–6, maximum 15)]. In the remaining 52 patients in whom SRCC was detected at least once [median follow-up 36 months (IQR: 18-61, maximum 149); median 5 endoscopies (IQR: 3-8, maximum 14)]. The median age at first detection of positive foci during endoscopy surveillance is 32.5 y (25-47 y, range 19-80).

The endoscopic findings appeared largely stable over time, with fluctuations rather than consistent progression (Figure 4). When random and targeted biopsies were presented separately, similar patterns were observed (Figure S3). Quantitative analysis with mixed-effects negative binomial regression confirmed the absence of a significant longitudinal trend in SRCC detection. The incidence rate ratio (IRR) for the number of positive foci in surveillance endoscopy did not increase over time (1.028 per six months, *P* = 0.177), across the entire surveillance cohort. These findings suggest that disease progression in HDGC is generally slow, with endoscopic findings characterized by temporal variability rather than steady progression.

**Figure 4.**
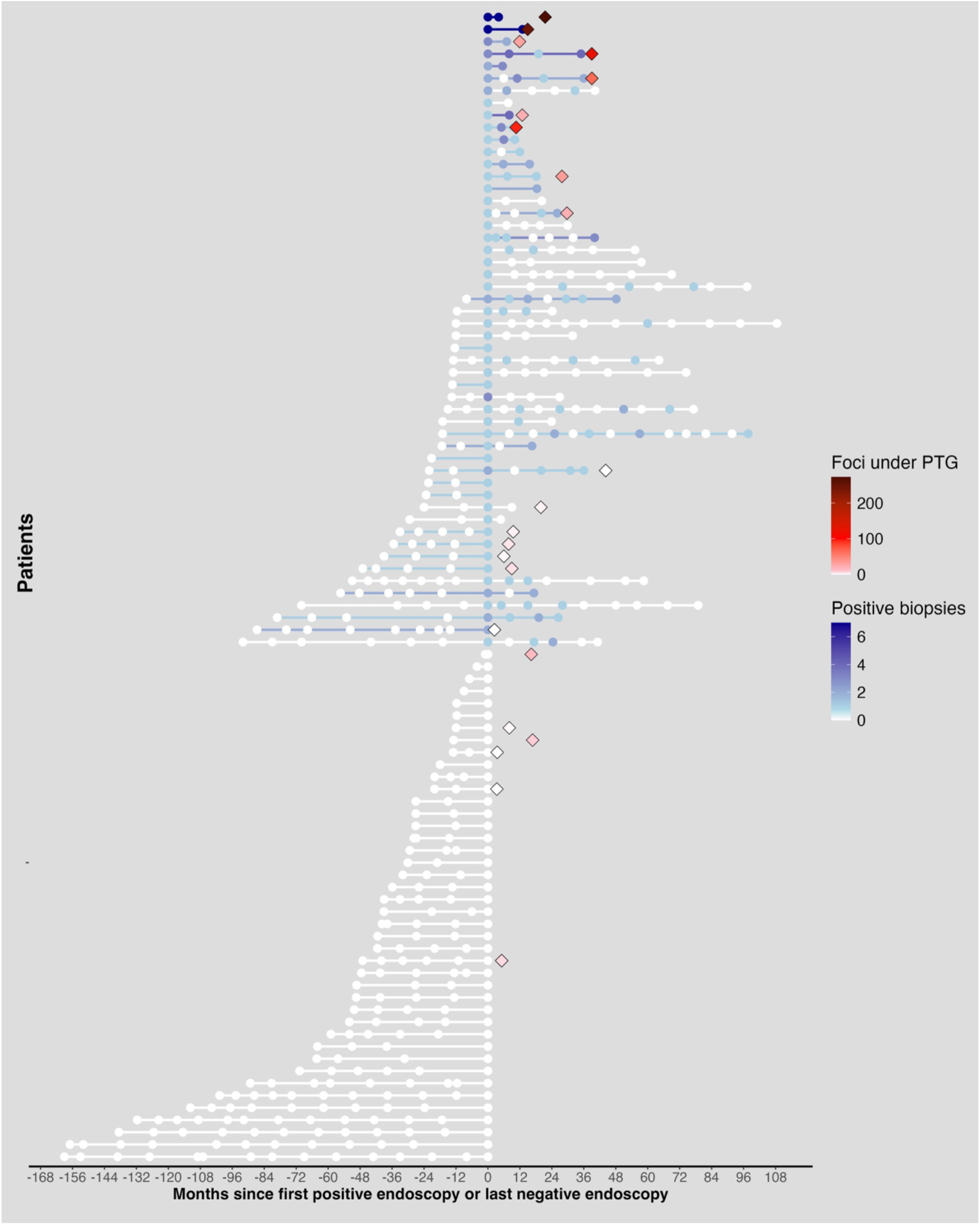
Endoscopic findings in *CDH1-GPV* carriers before and after signet ring cell carcinoma (SRCC) detected during surveillance endoscopy. Each row represents one HDGC individual who underwent endoscopic surveillance followed by PTG, and the lines were colored by the number of positive biopsies in the last endoscopy. Circles denote endoscopic procedures, colored by the number of positive biopsies (both random and targeted), with separated counts shown in Figure S3, while diamonds represent PTG specimens, colored by the total number of SRCC foci identified in gastrectomy specimens. The most recent endoscopies were excluded for one patients (the top case in the figure), as no biopsies were taken during their final procedure prior to PTG. A few patients without positive endoscopic biopsies underwent PTG based on their personal willingness. Endoscopy follow-up ended either at the time of prophylactic total gastrectomy (PTG) or by February 2024. Endoscopic findings remained largely stable over time, showing fluctuations rather than consistent progression.

### Endoscopic findings across various surveillance intervals

Among 52 patients in whom SRCC was detected in at least one endoscopy, 37 had 1–2 positive biopsies at their first positive examination and subsequently underwent at least one further surveillance procedure. The majority of subsequent surveillance procedures demonstrated stable findings: 62% at 6-month intervals, 65% at 12 months, and 66% at 24 months. An increase in endoscopically detected SRCC was infrequent, occurring in only 3% to 6% of cases across all intervals (Figure 5). A third of these patients (31%–32%) exhibited decrease, highlighting some degree of variability in biopsy outcomes over time. There was no progression to advanced cancer in patients with signet ring cell carcinoma foci on biopsies, either random or target from pale areas with no worrying features. Nevertheless, 19% (7/37) of these patients received PTG either based on physician advice due to increased number of SRCC foci on endoscopic biopsies (n=2) or due to personal preference (n=5).

**Figure 5.**
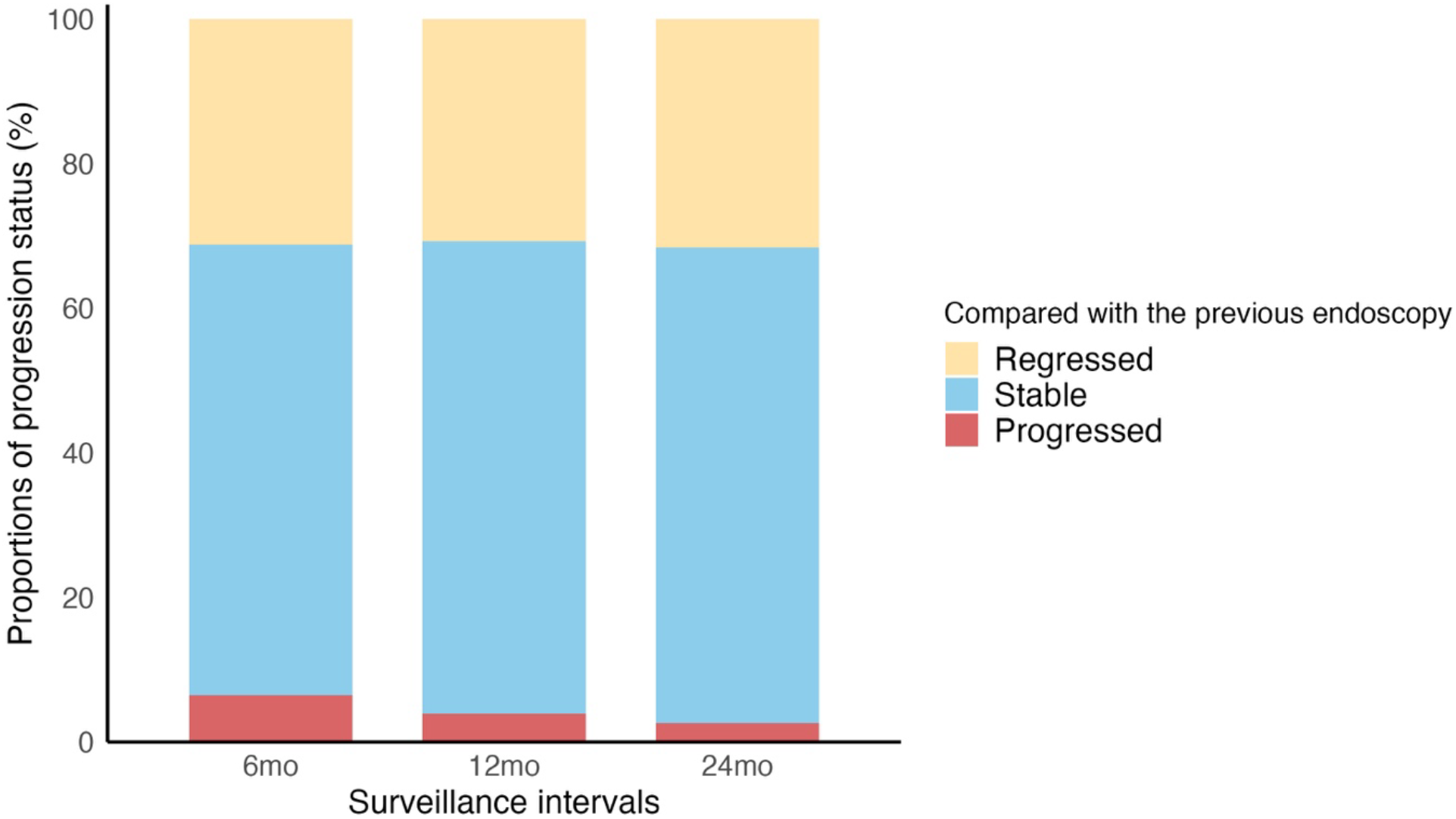
Simulated surveillance outcomes at 6-, 12-, and 24-month intervals. To evaluate the effectiveness of different surveillance intervals for low-risk positive patients had 1–2 positive biopsies at their first positive examination. Surveillance intervals were categorized into 6-, 12-, and 24-month groups based on the time elapsed between consecutive endoscopic procedures. For the 12- and 24-month interval groups, intermediate endoscopy records were masked to simulate true annual or biennial surveillance schedules. No statistically significant differences in surveillance outcomes were observed among the three groups.

## Discussion

This study demonstrates that in patients with HDGC it is possible to estimate the burden of early SRCC based on the integration of histopathology data from random and targeted biopsies. In addition, in a well-characterised longitudinal cohort of patients with no endoscopic features of advanced cancer, we demonstrate that the increase in the burden of endoscopic early cancer lesions is an infrequent event.

Given the aggressive nature of diffuse type gastric cancer and challenges related to endoscopic detection and risk stratification, PTG has been for decades the mainstay of treatment. This has led to the recommendation of surgery upon detection of SRCC on endoscopic biopsies..^10-12^ However findings challenge a blanket policy of major surgery as tretment of indolent and scarce foci of SRCC and confirm safety of endoscopic surveillance in such cases. Several case series and two larger longitudinal cohort studies have in fact demonstrated that progression to advanced cancer on surveillance is a rare event.^6,7,13^ Given the lack of outcomes in terms of progression to Stage >1 cancer, it is difficult to develop prediction models for progression to advanced cancer. For this reason, we focused in this study on models to predict low burden of early SRCC, in the assumption that individuals with low SRCC burden are at low progression risk. Although imperfect, this strategy reflects the current knowledge of cancer risk in other gastrointestinal conditions that predispose to cancer like Barrett’s oesophagus, gastric atrophy and ulcerative colitis, where the topographical extent of the disease correlates with the risk of cancer.^14-16^

Our data show that the number of SRCC foci on both random and targeted biopsies is the strongest predictor of early cancer burden and that the combination of the two types of biopsies achieves the best diagnostic accuracy. It has been debated whether mapping biopsies are useful in the monitoring of these individuals, with the argument that it is clinically insignificant to detect indolent microscopic foci of disease, given that around 90% of *CDH1-PV* carriers have microscopic foci detected in PTG specimens.^17^ Our data are supportive of the routine use random biopsies to provide the most comprehensive information for determining the best management strategies.

Current guidelines recommend that individuals with positive biopsy findings, who decline PTG, be seen for repeat OGD in 6 months.^4^ Our data not only demonstrate the safety of endoscopic monitoring even in the presence of targeted or random biopsies positive for SRCC, within the context of informed patient choice and multidisciplinary review, but also suggest current intervals for surveillance are too short especially in low-burden patients and early follow up unlikely to provide clinically useful information. In particular, our data indicate that a negative baseline endoscopy based on full mapping biopsies and high quality endoscopy with electronic chromoendoscopy and magnification is a strong predictor of low burden of disease meaning that follow up could be extended to in expert centres. Also, the findings suggest that a six month follow up is not necessary in the presence of a small number of biopsies positive for SRCC and in the absence of worrying endoscopic features (elevated or depressed lesions, ulcers or thickened folds).^17^ Based on the data, we believe it is safe and sensible for patients with low endoscopic cancer burden to be followed yearly rather 6-monthly. We propose in Figure 6 a modified surveillance protocol which is supported by evidence of safety in the current study. We envisage that future studies might provide new data to allow further adjustment of the intervals.

**Figure 6.**
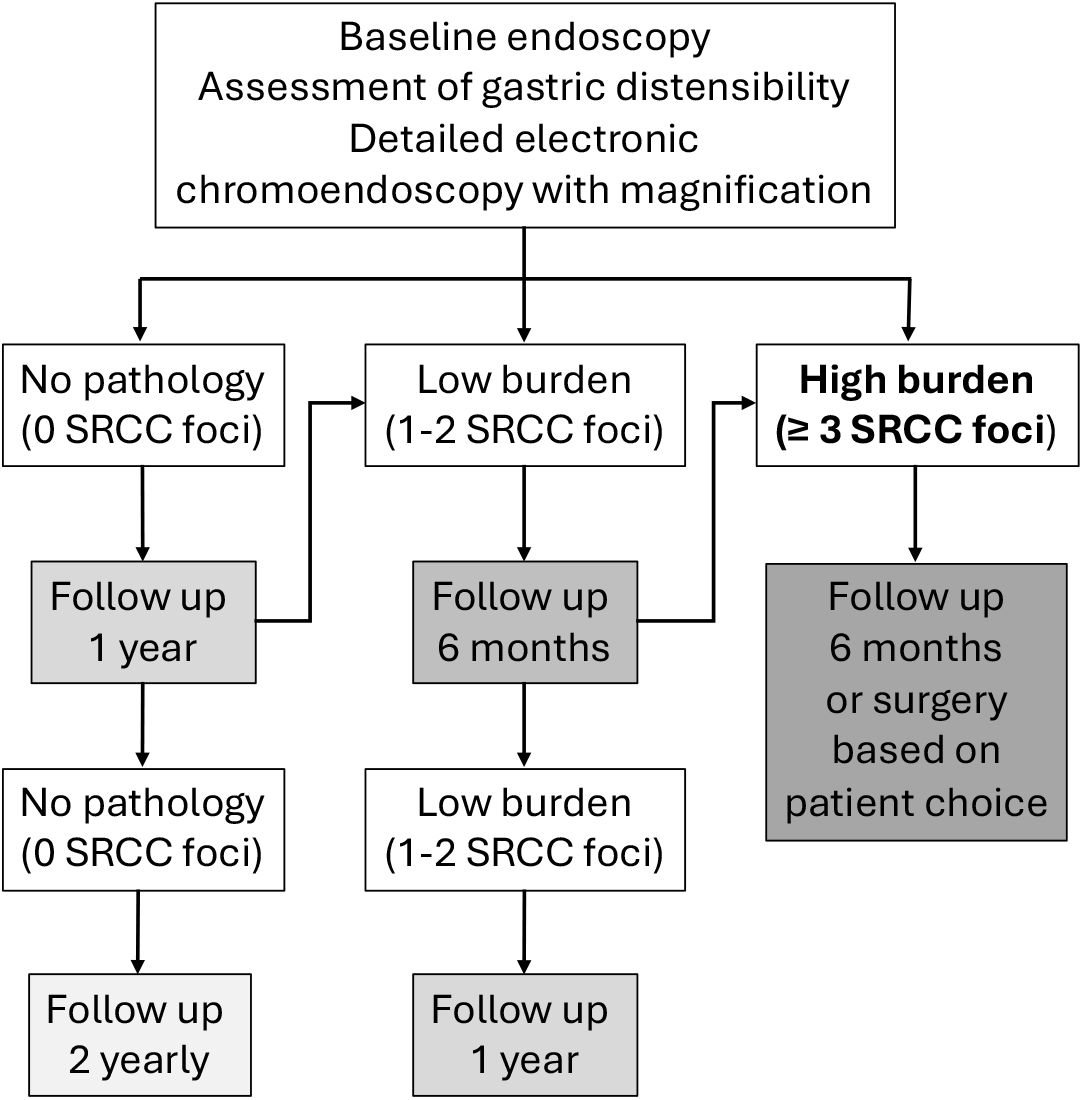
A modified surveillance protocol for hereditary diffuse gastric cancer individuals with *CDH1* germline pathogenic variation, supported by evidence of safety from the study.

Taken together our findings suggest that SRCC progression is generally uncommon under surveillance, and low endoscopic SRCC burden correlates with scarce early neoplastic gastric disease.

## Data Availability

All data produced in the present study are available upon reasonable request to the authors

## Author contribution

LW collected and analysed the data and wrote the manuscript. HJL collected the data and contributed to analysis and design. NK and SS assisted data collection. MO’D directed histopathological assessment of hereditary diffuse gastric cancer samples. MT provided input on genetic data. RON performed PTG and provided data. RCF is chief investigator on the Familial Gastric Cancer Registry study, conceived the biopsy protocol, provided guidance on the project. MdP conceived the project, performed endoscopic procedures, conceived the endoscopic diagnostic criteria, and wrote the manuscript. The corresponding author had final responsibility for the decision to submit the manuscript for publication. All authors reviewed the manuscript before submission.

---

**Figure S1.**
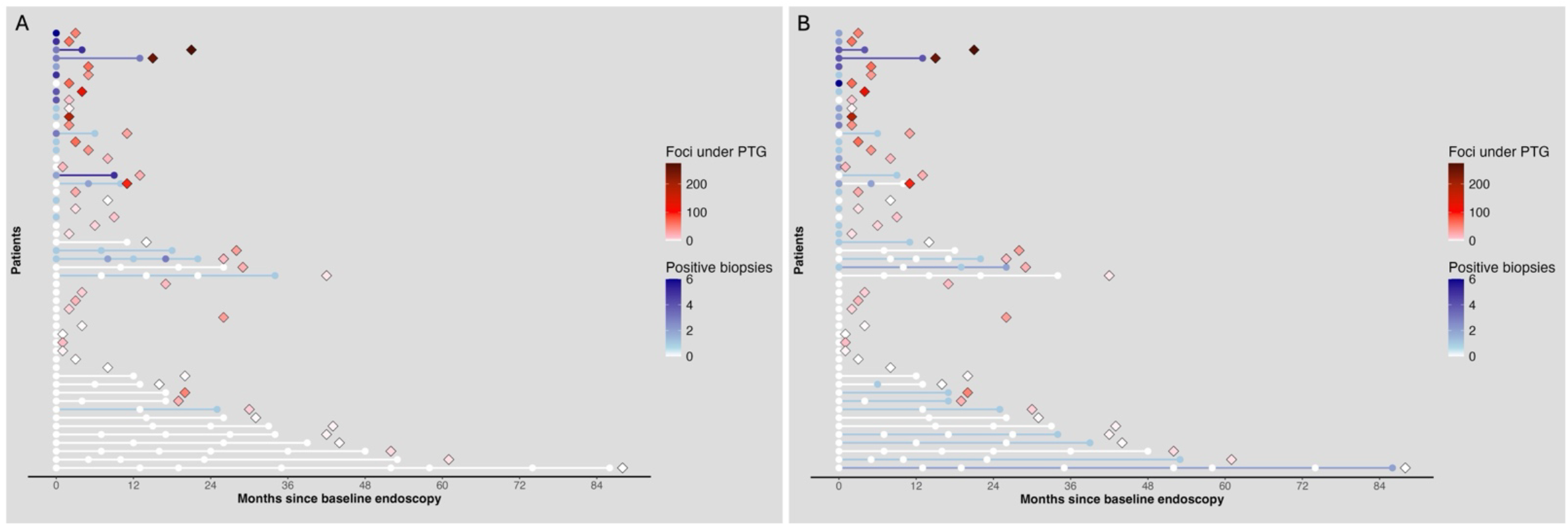
Impact of biopsy type on detecting high-burden cases. Heatmap showing which patients would be classified as “positive” at baseline based on biopsy type: targeted only (A), or random only (B). A number of patients with high SRCC burden (as confirmed in PTG) would have been missed if only one biopsy method was used, supporting the combined strategy.

**Figure S2.**
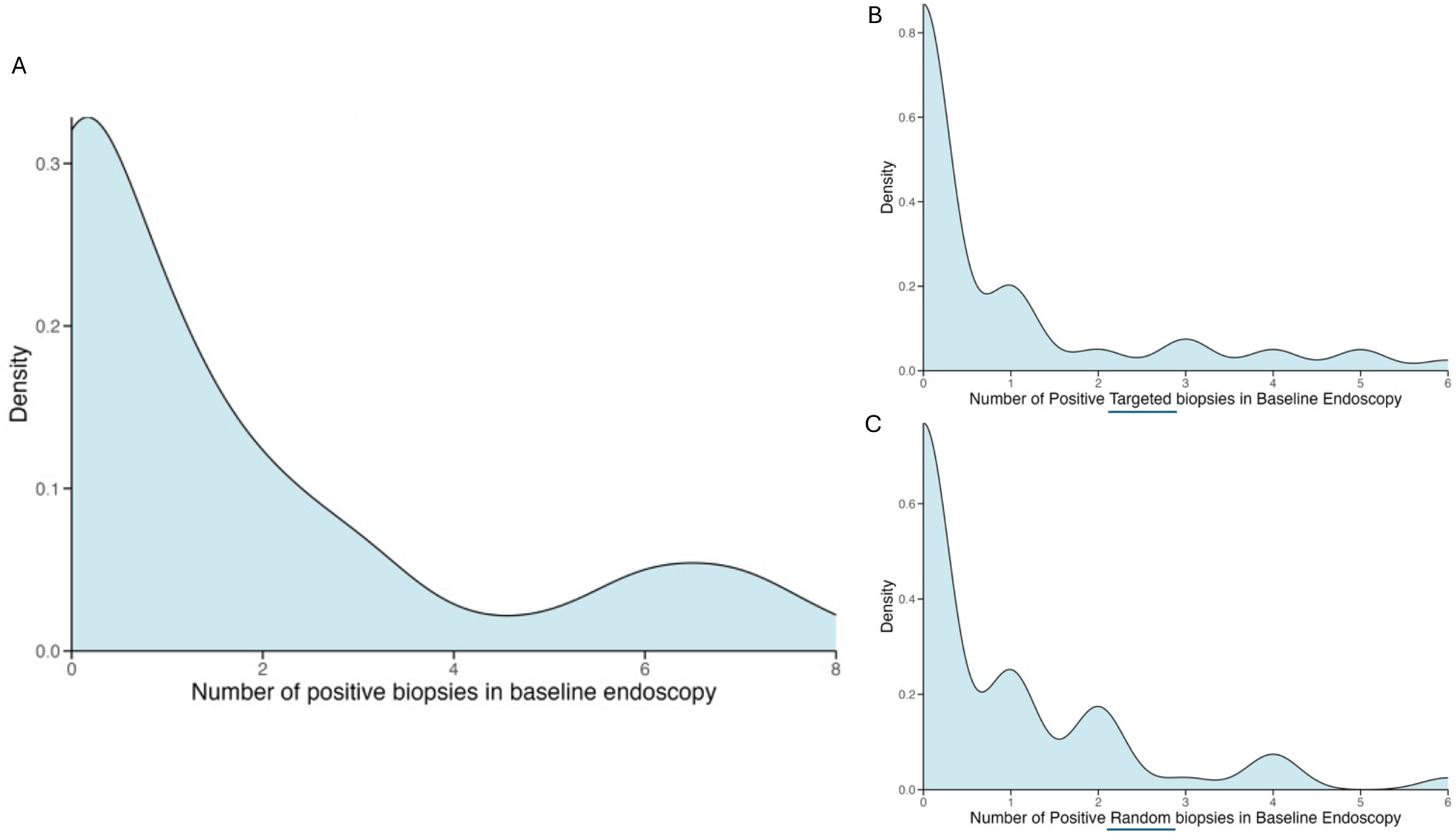
Density plots illustrating the right-skewed distribution of positive biopsies during surveillance endoscopy: (A) all biopsies, (B) targeted biopsies, and (C) random biopsies.

**Figure S3.**
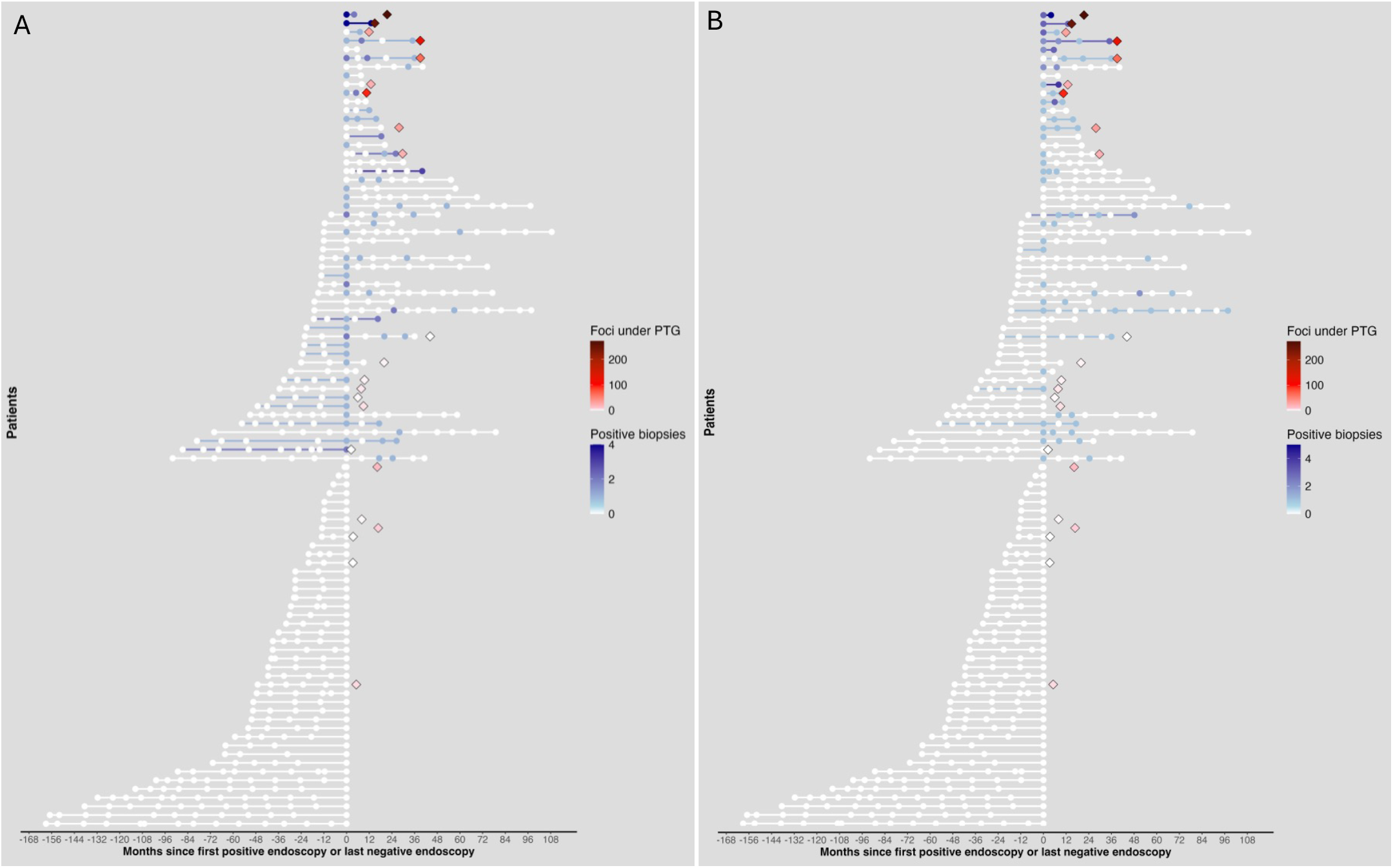
The number of positive random (A) and targeted (B) biopsies during surveillance endoscopy. The endoscopic findings appeared largely stable over time, with fluctuations rather than consistent progression. Each row represents one HDGC individual who underwent endoscopic surveillance followed by PTG, and the lines were colored by the number of positive biopsies in the last endoscopy. Circles denote endoscopic procedures, colored by the numbers the positive random (A) or targeted (B) biopsies. Patient order is consistent with Figure 2. Diamonds represent PTG specimens, colored by the total number of SRCC foci identified in gastrectomy specimens. The most recent endoscopies were excluded for one patients (the top case in the figure), as no biopsies were taken during their final procedure prior to PTG. A few patients without positive endoscopic biopsies underwent PTG based on their personal willingness. Endoscopy follow-up ended either at the time of prophylactic total gastrectomy (PTG) or by February 2024.

